# Analysis of the Temporal Dynamics of Dengue, Zika and Chikungunya in Ecuador: Emergency Patterns and Associations Epidemiological (1988-2024)

**DOI:** 10.1101/2025.04.25.25326457

**Authors:** José Daniel Sánchez, Carolina Álvarez Ramírez, Emilio Cevallos Carrillo, Juan Arias Salazar, Cesar Barros Cevallos

## Abstract

**Summary:** This study analyzes the temporal dynamics and possible epidemiological interactions of three arboviruses transmitted by Aedes mosquitoes in Ecuador: dengue (DENV), Zika (ZIKV) and chikungunya (CHIKV), during the period 1988-2024.

Using national epidemiological data, we examined incidence patterns, potential associations between these diseases, and their relationship with temporal factors. The results reveal cyclical patterns of dengue, with established endemicity and notable epidemic peaks (1994, 2000, 2015, 2024), with transmission progressively expanding to ecologically diverse regions. Zika and chikungunya showed more recent explosive introductions, with significant outbreaks concentrated in 2015–2017, followed by a dramatic decline. The analysis suggests a complex epidemiological interaction, where the emergence of one arbovirus seems to coincide with changes in the incidence of another. Population immunity, vector dynamics influenced by socioecological and climatic factors, and public health responses likely modulate these patterns. This study underscores the need for integrated surveillance and adaptive control strategies for these arboviruses in Ecuador.

## 1. Introduction

Mosquito-borne arboviruses, particularly dengue, Zika, and chikun-gunya, represent a growing public health challenge in tropical and subtropical regions, including Ecuador (Loor Frank et al., 2023, oor Frank et al., 2023). These viruses, transmitted primarily by mosquitoes of the genus Aedes (predominantly Aedes aegypti in Ecuadorian urban and peri-urban environments), share common vectors and initial clinical symptoms, complicating the differential diagnosis and healthcare response (Talbot et al., 2021, albot et al., 2021; Loor Frank et al., 2023, oor Frank et al., 2023).

Ecuador, with its ecological diversity, favorable climatic conditions (including the influence of phenomena such as El Niño/La Niña), variable socioeconomic factors, and the established presence of the vector, offers an environment conducive to the endemic and epidemic transmission of these diseases (Katzelnick et al., 2024; Sippy et al., 2019; Sippy et al., 2019).

The epidemiological history of dengue in Ecuador is extensive, with endemic transmission in several regions, especially on the coast, since the end of the 20th century (Katzelnick et al., 2024, Atzelnick et al., 2024). However, the almost simultaneous introduction of chikungunya (2014-2015) and Zika (2015-2016) in the Americas, including Ecuador, created a new epidemiological scenario characterized by the co-circulation of multiple arboviruses (Ochoa Asanza & Jerson Amado, 2018, Choa Asanza & Jerson Amado, 2018; Katzelnick et al., 2024, Atzelnick et al., 2024).

This study aims to analyze the temporal patterns of dengue (including severe dengue), Zika, and chikungunya incidence reported in Ecuador during the period 1988–2024. We seek to identify trends, epidemic cycles, and the sequence of emergence of these viruses, and to explore possible associations or temporal shifts in their occurrence patterns that may suggest epidemiological interactions.

## 2. Methods

### 2.1. Data Sources

Aggregated epidemiological surveillance data on reported cases of dengue, severe dengue, Zika, and chikungunya in Ecuador for the period 1988–2024 were used. These data, collected and consolidated by the Ministry of Public Health (MSP) of Ecuador through its national surveillance system, include the annual number of reported cases. Annual incidence rates per 100,000 inhabitants were calculated using national population estimates and the GIDEON database for the country (https://app.gideononline.com/start).

### 2.2. Data Analysis

A descriptive analysis of the time series of reported cases and incidence rates was performed for each arbovirus. Temporal trend graphs were generated to visualize incidence patterns throughout the study period. Years with epidemic peaks (defined as substantial increases above reference levels) and periods of low incidence were identified for each disease.

The temporal co-occurrence of epidemics was qualitatively assessed by examining the sequence of introduction and the periods of overlap or possible displacement in their incidence peaks. The analysis is contextualized with findings from published studies on the epidemiology, risk factors, and vector dynamics of arboviruses in Ecuador (Katzelnick et al., 2024; Atzelnick et al., 2024; Sippy et al., 2019; Sippy et al., 2019; Talbot et al., 2021; Talbot et al., 2021).

**Figure 1:**
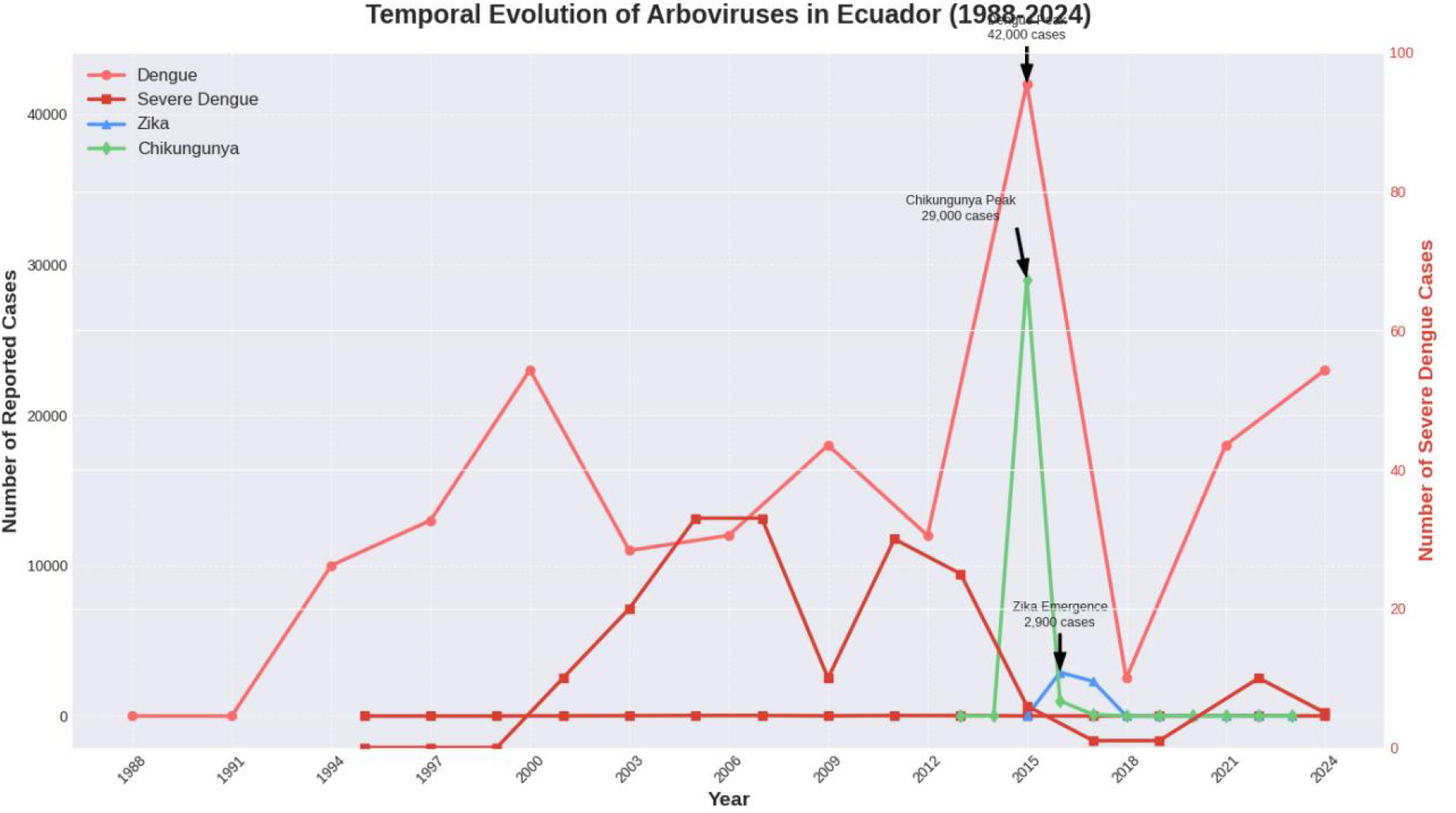
This title indicates that the graph shows the evolution or change over time (from 1988 to 2024) of the number of arbovirus cases reported in Ecuador.

### 2.3. Ethical Considerations

This study is based on aggregated and anonymized public health surveillance data and therefore did not require specific ethical approval.

## 3. Results

### 3.1. Temporal Trends by Disease

#### 3.1.1. Dengue

Dengue has shown a persistent endemic presence in Ecuador since at least the early 1990s, with documented transmission intensifying around 1988 and major epidemic peaks recorded in:

- 1994: ÿ10,000 cases (ÿ80 per 100,000 inhab.)
- 1997: ÿ13,000 cases (ÿ100 per 100,000 inhab.)
- 2000: ÿ23,000 cases (ÿ150 per 100,000 inhab.)
- 2009: ÿ18,000 cases (ÿ120 per 100,000 inhab.)
- 2015: Historic peak with ÿ42,000 cases (ÿ300 per 100,000 inhab.)
- 2024: New significant peak with ÿ23,000 cases (ÿ150 per 100,000 inhab.)

This pattern suggests recurrent epidemic cycles, with a geographic expansion observed from coastal provinces to inland and Amazonian regions in recent years.

two decades (Katzelnick et al., 2024, atzelnick et al., 2024). Coastal provinces such as Guayas, Manabí, and Esmeraldas consistently report high loads (Loor Frank et al., 2023, oor Frank et al., 2023).

#### 3.1.2 Severe Dengue

Cases of severe dengue (formerly dengue hemorrhagic fever) began to be reported. consistently from 2001 onwards. Moderate peaks were observed in:

- 2005 and 2007: ÿ33 cases (ÿ2.25 per 100,000)
- 2011: ÿ30 cases (ÿ2 per 100,000)
- 2013: ÿ25 cases (ÿ1.75 per 100,000)

Notably, in recent years, despite high numbers of dengue cases (e.g., 2024), reported cases of severe dengue have decreased considerably, with only ÿ10 cases in 2022 (ÿ0.5 per 100,000) and ÿ5 cases in 2024 (ÿ0.25 per 100,000).

#### 3.1.3. Zika

The Zika virus emerged epidemiologically in Ecuador in 2016, causing a significant outbreak:

- 2016: ÿ2,900 cases (ÿ25 per 100,000 inhab.)
- 2017: ÿ2,300 cases (ÿ20 per 100,000 inhab.)

From 2018 to 2023, reported cases were extremely sporadic according to the data analyzed. The emergency was rapid and affected several provinces.

#### 3.1.4. Chikungunya

The Chikungunya virus emerged a year before Zika, in 2015, causing an even larger epidemic:

- 2015: ÿ29,000 cases (ÿ250 per 100,000 inhab.)
- 2016: Dramatic decrease to ÿ1,000 cases (ÿ10 per 100,000 inhab.)
- 2017: ÿ100 cases (ÿ1 per 100,000 inhab.)

Similar to Zika, reported cases of chikungunya have been virtually absent. from 2018 to 2023.

**Figure 2:**
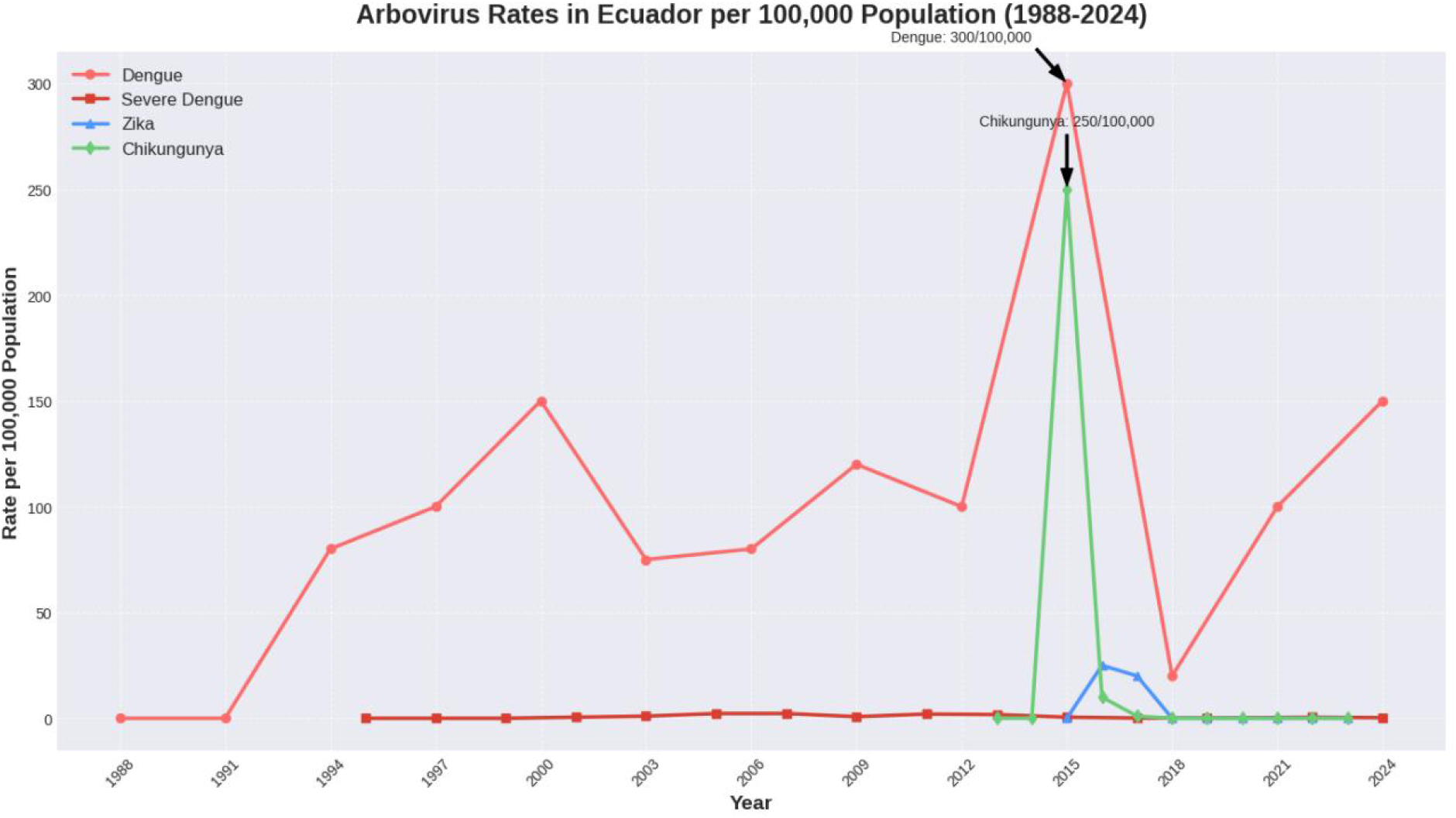
This graph shows the incidence rate (new cases per 100,000 inhabitants) of four arboviruses (Dengue, Severe Dengue, Zika and Chikungunya) in Ecuador during the period 1988-2024. Unlike the previous graph, which showed absolute numbers, this one normalizes cases according to population, allowing comparison of the intensity the impact of diseases over time.

### 3.2. Temporal Association Patterns between Arboviruses

1. Sequential Introduction: A clear sequence is observed in the epidemic emergence of these viruses: Dengue (endemic with peaks since the 90s), Chikungunya (epidemic in 2015) and Zika (epidemic in 2016).
2. Coincidence and Shift of Peaks: The year 2015 was exceptional, marking the largest dengue epidemic on record and the massive outbreak of chikungunya. The Zika emergency in 2016 coincided with a significant decline in both of dengue and chikungunya cases compared to their peaks this year former.
3. Post-Emergency Synchronous Decline: Both Zika and Chikungunya showed a drastic reduction in reported cases after 2017, while Dengue continued to circulate and caused a new epidemic peak in 2024.
4. Inverse Dengue/Severe Dengue Relationship: The downward trend in the notification of severe dengue in recent years contrasts with the persistence and recent peaks in overall dengue incidence.

## 4. Discussion

The temporal patterns observed in the incidence of dengue, Zika and chikungunya in Ecuador between 1988 and 2024 suggest a complex epidemiology, probably influenced by caused by a combination of immunological, virological, vector, and environmental factors and socioeconomic, as well as public health interventions.

### 4.1. Possible Viral Interactions

The sequence of introduction and the observed movement patterns, particularly the decline in dengue and chikungunya coinciding with the rise of Zika in 2016, support the hypothesis of interactions between these arboviruses. Potential mechanisms include:

#### 4.1.1. Cross-immunity

Previous infection with a flavivirus (such as dengue) may confer partial immunity or modify the response to subsequent infection with another flavivirus (Zika), although the evidence is complex and the effects may vary. Interactions between flaviviruses (DENV, ZIKV) and alphaviruses (CHIKV) are less likely at the direct immunological level, but could occur at the level of host response or competition for resources cell phones.

#### 4.1.2. Vectorial Competence

Aedes mosquitoes can be infected with and transmit all three viruses. Infection with a A mosquito with one virus could reduce its susceptibility or ability to transmit another subsequently acquired virus (intrinsic competence). Furthermore, intensified vector control interventions during large outbreaks (such as those of CHIKV and ZIKV in 2015-2016) could have temporarily reduced mosquito populations, impacting the transmission of all viruses that share the vector (Katzelnick et al., 2024, atzelnick et al., 2024).

### 4.2. Dengue Dynamics

The endemic persistence and epidemic cycles of dengue (approximately every 6-9 years in this analysis) are characteristic of this disease and probably reflect the complex interaction between population immunity (which declines over time and is serotype-specific), the circulation and introduction of different serotypes of the DENV virus (1-4), and environmental factors.

Studies in Ecuador have correlated dengue transmission with climatic factors such as temperature and precipitation (Sippy et al., 2019, Sippy et al., 2019), and socio-ecological factors such as urbanization, access to water, waste management and housing conditions, which influence the density of the Aedes vector (Katzelnick et al., 2024, Atzelnick et al., 2024; Talbot et al., 2021, Talbot et al., 2021). The expansion observed geographic (Katzelnick et al., 2024, atzelnick et al., 2024) underlines the growing adaptability of the vector and/or virus.

### 4.3. Emergence and Decline of Zika and Chikungunya

The “burst and disappearance” pattern of Zika and Chikungunya is common following the introduction of a new pathogen into a fully susceptible population. The rapid Initial spread probably depleted a large proportion of the susceptible population, leading to temporary herd immunity that hampered sustained transmission.

However, the virtual absence of reported cases in subsequent years could also be influenced by changes in the intensity of ZIKV and CHIKV-specific surveillance, and the difficulty in differential diagnosis with dengue and other febrile illnesses (Ochoa Asanza & Jerson Amado, 2018, Ochoa Asanza & Jerson Amado, 2018). These viruses may continue to circulate at low levels or may reemerge if population immunity wanes.

### 4.4. Reduction of Severe Dengue

The apparent reduction in the proportion of severe dengue cases relative to total dengue cases is a positive finding but requires further investigation. It could be due to:

- Improvements in clinical management and early detection
- Changes in the virulence of the predominant circulating serotypes/genotypes
- Modulation of severity due to prior exposure to other flaviviruses (such as Zika) or different serotypes of dengue

The lack of systematic serotypic data is a key limitation in elucidating this.

## 5. Conclusions

Analysis of the temporal dynamics of dengue, Zika, and chikungunya in Ecuador (1988–2024) reveals distinct patterns: dengue persists as an endemic-epidemic problem with recurrent cycles and geographic expansion, while Zika and chikungunya caused intense but transient epidemics after their recent introduction. Co-occurrence and temporal displacement suggest possible epidemiological interactions between these arboviruses, which share the same primary vector.

These findings reinforce the critical need for robust, integrated epidemiological and entomological surveillance systems in Ecuador. Control strategies must be adaptive, considering the possibility of co-circulation and potential interactions between arboviruses, and must address the local socio-ecological determinants that favor the proliferation of the Aedes vector.

## 6. Limitations and Recommendations

This study is based on aggregated reporting data, which may be subject to underreporting and diagnostic bias, especially for Zika and Chikungunya after their initial epidemic phases and due to their clinical similarity to dengue (Ochoa Asanza & Jerson Amado, 2018, Ochoa Asanza & Jerson Amado, 2018). Changes in case definitions or diagnostic capacity over more than three decades could also influence the observed trends.

The lack of regionally stratified data and the absence of information on circulating DENV serotypes/genotypes limit the depth of analysis of interactions and severity patterns. It is recommended:

1. Strengthen integrated surveillance of arboviral diseases, including timely reporting, laboratory confirmation, and molecular characterization. (serotypes/genotypes) of circulating viruses.
2. Conduct longitudinal seroprevalence studies to better understand population immunity against DENV, ZIKV and CHIKV and assess the magnitude of the potential cross-immunity.
3. Intensify entomological surveillance and research on local socio-ecological factors that determine the density and competence of the Aedes vector (Talbot et al., 2021, albot et al., 2021; Katzelnick et al., 2024, atzelnick et al., 2024).
4. Specifically investigate the biological and epidemiological mechanisms of the interactions between these arboviruses in the Ecuadorian context.
5. Use these improved data to develop predictive models that can inform vector control and public health strategies more effectively.

## Data Availability

The data used in the study comes from the Ministry of Public Health (MSP) of Ecuador through its national surveillance system, and the GIDEON database (https://app.gideononline.com/start)

## Notes

### Competing Interest Statement

The authors have declared no competing interest.

### Funding Statement

funding statement is not mentioned in the provided document

